# Hormonal contraception and SSRI’s influence on oxytocin and cortisol reactivity to stress in patients with borderline personality disorder

**DOI:** 10.64898/2025.12.18.25342546

**Authors:** M. Teixeira de Almeida, L. Quattrocchi, N Perroud, T. Aboulafia-Brakha

## Abstract

**Background:** Borderline personality disorder (BPD) involves emotional instability and stress sensitivity linked to oxytocinergic and HPA-axis dysregulation. This study examined oxytocin and cortisol responses to acute psychosocial stress and the modulatory effects of SSRIs and hormonal contraception.

**Methods:** 93 female participants (45 with BPD with/without SSRIs and 48 healthy controls) underwent the Trier Social Stress Test. Linear and generalized linear mixed-effects models were applied to assess time, group, and hormonal contraception effects, and their interactions.

**Results:** During the stress task, both BPD groups reported significantly higher subjective anxiety and anger than controls. All participants showed increased salivary oxytocin (OXT) during stress anticipation, but post-task trajectories diverged: BPD participants without SSRIs exhibited a sharp OXT decline, whereas those on SSRIs mirrored the stable trajectory of controls. Hormonal contraception reversed the OXT decline in untreated BPD participants, resulting in a progressive increase during recovery. Cortisol (CORT) analyses revealed hyporeactivity in BPD participants without SSRIs, a pattern unaffected by hormonal contraception. A significant three-way interaction indicated that higher OXT levels were associated with higher CORT concentrations during late recovery specifically in the BPD SSRI group, a relationship that was marginal in untreated patients and absent in controls.

**Conclusions:** Our findings confirm that neuroendocrine dysregulation in BPD is context-dependent and sensitive to pharmacological modulation. The ability of SSRIs and hormonal contraception to influence stress-response patterns, despite limited efficacy on core symptoms, highlights the importance of controlling for medication and hormonal status in future BPD biomarker research.

## 1. Introduction

Borderline personality disorder (BPD) is a psychiatric condition characterized by emotional lability, intense fears of abandonment, and unstable interpersonal relationships (American Psychiatric Association, 2013). It has an estimated prevalence of 1–2% in the general population, with comparable distribution between men and women, although women are more frequently represented in clinical samples (Bozzatello et al., 2024). The disorder is frequently associated with high rates of psychiatric comorbidities, including major depressive disorder, post-traumatic stress disorder (PTSD), social anxiety, and other anxiety disorders, thereby contributing to diagnostic and therapeutic challenges (Leichsenring et al., 2023; Biskin & Paris, 2013). Importantly, stressful contexts often trigger or intensify core BPD symptoms, highlighting the relevance of investigating stress reactivity in this disorder (Glaser et al., 2008).

The stress response is mediated by a coordinated hormonal cascade, the most well-known being the hypothalamic–pituitary–adrenal (HPA) axis, which culminates in cortisol (CORT) release from the adrenal cortex (Herman et al., 2016). Growing evidence indicates that oxytocin (OXT) plays also an essential role in stress regulation. OXT is synthesized in the hypothalamus and released via the posterior pituitary, and is traditionally associated with attachment, social bonding, and reproductive processes such as parturition and lactation (Baribeau & Anagnostou, 2015; Alves et al., 2015; DeLaMare, 2023). Beyond its well-established functions in reproduction and social affiliation, OXT modulates emotional and physiological responses to stress, exerting anxiolytic effects in psychosocial contexts (Amico et al., 2004; Donadon et al., 2018). In healthy individuals, OXT levels typically rise early during stress exposure. This increase appears to down-regulate the subsequent CORT peak during the recovery period (de Jong et al., 2015; Bernhard et al., 2018; Alley et al., 2019), suggesting that OXT plays a regulatory role in stress response.

In BPD, this hormonal cascade seems to be disrupted, with either heightened or reduced basal (resting) CORT levels (Lieb et al., 2004; Thomas et al., 2019) and blunted or delayed CORT release during stress, as observed during the Trier Social Stress Test (TSST), a standardized laboratory paradigm known to reliably elicit robust psychological and physiological stress responses (Nater et al., 2010; Walter et al., 2008). Such attenuated reactivity may reflect chronic stress exposure, altered HPA-axis feedback sensitivity, or comorbid conditions such as PTSD or depression. Regarding OXT, both basal and reactive (stress-induced) hormonal profiles appear altered, but findings are inconsistent. Several studies report reduced basal plasma or serum OXT in BPD compared with healthy controls (Bertsch et al., 2013; Carrasco et al., 2020; Ebert et al., 2018), whereas others report no group differences (Bomann et al., 2017; Bonfig et al., 2022). In one of our recent work we observed no significant differences between BPD and control participants during basal circadian assessment and both groups showed a consistent pattern of increase from awakening to noon (Teixeira de Almeida et al., 2025). Importantly, there seems to be a hight heterogeneity within BPD with reduced OXT levels are particularly pronounced in individuals with disorganized attachment (Jobst et al., 2016). In contrast, reactive OXT responses show more convergent alterations. Multiple studies report blunted or decreased OXT release during social exclusion or interpersonal closeness paradigms in BPD, indicating an impaired OXT response to social stress (Jobst et al., 2016; Bonfig et al., 2022).

Several factors, such as medication and hormonal contraception, can also affect CORT and OXT levels. A study by Uvnäs-Moberg et al. (1999) showed that selective serotonin reuptake inhibitors (SSRIs) induced higher OXT plasma levels. CORT levels have also been shown to be affected by SSRIs, by increasing basal CORT levels in the evening (Manthey et al., 2011). Hormonal contraception also disrupts OXT and CORT levels by increasing them among oral contraceptive users (Garforth et al., 2020; Hertel et al., 2020). Therefore, when studying OXT and CORT responses, it is crucial to account for medication and contraceptive use, as these factors may impact results.

Despite evidence implicating dysregulation of both CORT and OXT systems in BPD, no studied have examined how their interplay operates under acute psychosocial stress in this population. Hence, the present study aims to investigate stress reactivity in individuals with BPD, specifically focusing on this hormonal interplay and the potential influence of SSRI and hormonal contraception on the physiological response

## 2. Methods

### 2.1 Participants

All participants provided written informed consent, and the study was approved by the local ethics committee (Swisethics ID: 2022-00067) and registered on clinicaltrial.gov (NCT05357521). A total of 128 female participants were recruited, of whom 93 completed the full protocol. Of these, 45 had a BPD diagnosis according to the DSM-5 criteria and were recruited through the Emotion Regulation Unit of the psychiatry department at Geneva University Hospitals; 48 were healthy controls, who met no more than 2 DSM-5 BPD criteria, and were recruited via advertisement. Clinical eligibility was verified using the BPD module of the Semi-Structured Clinical Interview for DSM-5 Axis II (SCID-5-PD BPD) (Ekselius et al.,1994) and the Mini International Neuropsychiatric Interview for DSM-5 disorders (MINI). The latter assesses for DSM-5 axis I major psychiatric conditions (Lecrubier et al., 1997).

Participants were also required to be between 18 and 35 years of age and have no history of neurological or neurohormonal disorders. General exclusion criteria included pregnancy, breastfeeding within six months prior to enrolment, the use of systemic corticosteroid medication, a body mass index (BMI) below 17, or a history of alcohol or drug addiction. Specific exclusion criteria for the BPD group included a diagnosis of psychosis (except for psychotic symptoms within BDP), a current manic episode, or the daily use of neuroleptics or non-SSRI/SNRI antidepressants. To control for the potential confounding effects of acute stress, participants who had experienced a major life stress event (e.g., death of a loved one, a break-up, or moving) within two months of inclusion were evaluated, and their enrolment was postponed if necessary.

### 2.2 Procedures

Our protocol consisted of three visits. An initial visit in which participants provided written informed consent and completed demographic, clinical, and baseline psychometric assessments. A second visit related to a naturalistic salivary sampling protocol conducted over a weekday in participants’ usual environment; for which procedures and results are described in Teixeira de Almeida et al. (2025). And finally, a third visit held one to four days later, in which participants performed an experimental stress task during which perceived emotional states and hormonal reactivity were assessed (the focus of this article). Of note, for participants not using hormonal contraception, sampling was scheduled during the luteal phase of the menstrual cycle.

### 2.3 Experimental stress task (Trier Social Stress Task-TSST)

The trier Social Stress Test (TSST) (Kirschbaum, Pirke, & Hellhammer, 1993) was administered according to standard procedures and mainly consisted of a resting period, followed by a 5-minutes prepared mock job interview and a 5-minutes surprise mental arithmetic task performed in front of two evaluators. One of the evaluators had no prior contact with the participant, to maintain social-evaluative novelty and increase stress induction reliability.

The experimental session started between 1pm and 1.30 pm to control for circadian variations of hormonal activity. Participants had their last meal no later than 12pm and, upon arrival at the laboratory, drunk a 20 cl natural apple juice containing 22 g of sugar to ensure stable sugar levels. They were then instructed to rest quietly for 20 minutes, while listening to a standardized playlist with classical music, without access to their phones or external communication. Baseline hormonal samples (as described in section 2.4) were then collected, followed by subsequent samples during the anticipation phase (5 minutes after instructions and right before the task), post-task (immediately after the TSST), and twice during recovery, respectively 15 and 50 min after the end of the task. . Self-reported questionnaires on state anger and stress/anxiety (section 2.4) were also completed at baseline (after rest), post-task and at 50 minutes recovery. The detailed procedure is illustrated in **Figure 1**.

**Figure 1.**
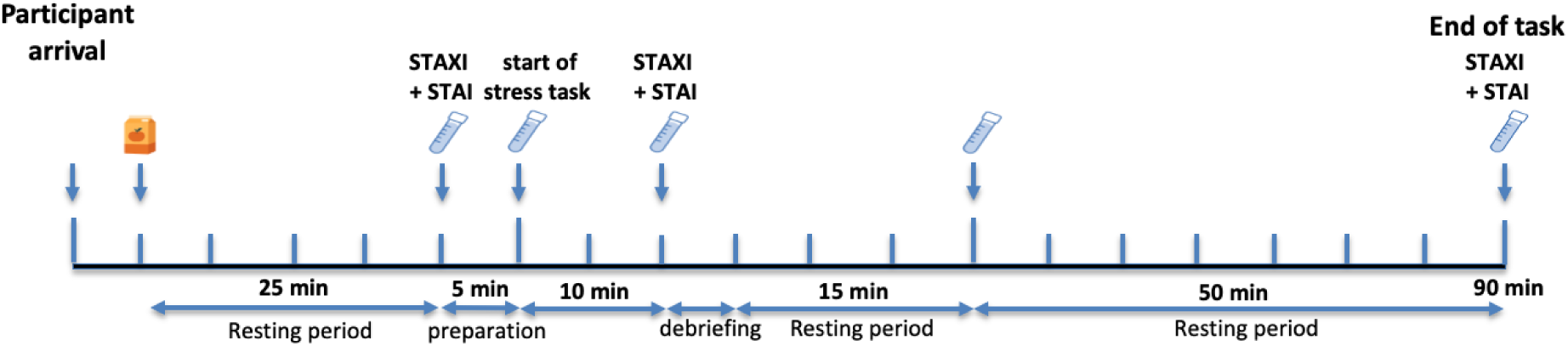
Schematical representation of the experimental session.

### 2.4 Measures

#### Demographic and clinical characteristics

We recorded biological age, Body Mass Index (BMI), years of education (from the start of primary school to the completion of the participant’s highest educational degree). We also recorded whether they practiced regular physical activity and if they were engaged in a stable romantic relationship (if applicable, the length of this relationship was recorded). Sleep duration was also calculated from bedtime sleep to wake-up time given by participants. Participants completed self-report questionnaires assessing depressive symptoms with the Beck Depression Inventory-BDI-II (Beck, et al., 1996)), anxiety traits with the State-Trait Anxiety Inventory-STAI (Spielberger et al., 1971)) and anger traits with the State-Trait Anger Expression Inventory- STAXI (Borteyrou et al., 2008; Spielberger et al. 1995). The Adverse Childhood Experiences (ACE) (Felitti et al., 1998) questionnaire and the Post-Traumatic Stress Disorder Checklist for DSM-5 (PCL-5) (Weathers et al., 2013) were used to record respectively exposure to childhood trauma before the age of 18 years old and to quantify post-traumatic stress disorder symptoms. Further, cognitive strategies used to regulate emotions after negative events were assessed with The Cognitive Emotion Regulation Questionnaire (CERQ) (Jermann et al., 2006) and adult attachment style with The Relationship Scales Questionnaire (RSQ) (Griffin & Bartholomew, 1994). The latter differentiates secure, fearful, dismissing, and preoccupied attachment styles.

#### Experimental task measures

##### Self-reported anger and stress

State anger and anxiety during the task were quantified with the state versions of the self-reported STAI (Spielberger et al. 1971) and STAXI (Borteyrou et al. 2008; Spielberger et al. 1995). These questionnaires were administered at three time-points: after the resting period (before the stress task), right after the stress task and at the end of the recovery period (50 minutes after the stress task). Higher scores indicate higher levels of state anger and stress.

##### Hormonal sample collection, processing and quantification

Saliva was obtained using Salivettes equipped with synthetic swabs (Sarstedt, Germany) at 5 different time points during the experimental task as described in *section 2.3*. Participants were instructed to gently chew the swab for a minimum of one minute to ensure sufficient sample volume. Samples were immediately stored at –20 °C. For extended storage, saliva was centrifuged and aliquots were placed at –80 °C until analysis. Each sample was divided into two aliquots, enabling parallel analysis of oxytocin and cortisol. All samples were shipped on dry ice to ensure they remained frozen throughout transport. Oxytocin, concentrations (pg/ml) were determined using radioimmunoassay (RIAgnosis, Sinzing, Germany) following established protocols described in prior publications (Aboulafia-Brakha et al., 2023; Bernhard et al., 2018; Jong et al., 2015; Teixeira de Almeida et al., 2025, Cazorla et al., 2025). Cortisol concentrations(µg/dl) were measured with a direct ELISA kit for human saliva (IBL Tecan). Analyses were conducted at the FCBG in Geneva, Switzerland. The method yielded an intra-assay variability of 4.3 % and an inter-assay variability of 13.2%.

### 2.5 Statistical Analyses

Data were analyzed using RStudio (R v4.2.2 / 4.4.1). Six participants (2 controls, 4 BPD) were excluded due to outlier values exceeding 1.5 × IQR based on the main dependent variables. Participants were divided into three groups (Control, BPD without SSRI, and BPD with SSRI) due to the potential influence of antidepressants on cortisol and oxytocin (Duman et al., 2004; Manthey et al., 2011). Comparisons between groups on demographic and clinical data were performed with a one-way ANOVA or Chi-square test, depending on the variable nature. Salivary oxytocin values followed a normal distribution and were analysed with linear mixed-effects models (LMM’s). Cortisol values were not normally distributed and were analysed with generalized linear mixed-effects models (GLMM) using a Gamma distribution and log link. Each model tested the main effects of group and time, followed by their group × time interaction. Random intercepts were included for participant ID to account for repeated measures. To investigate the interplay between cortisol and oxytocin, assessing for instance whether oxytocin levels predicted cortisol responses, we fitted an additional GLMM where cortisol was the dependent variable and oxytocin, group, and time were entered as interacting predictors. The use of hormonal contraception was inserted as a covariable in all models.

## 3. Results

### 3.1 Demographic and clinical data

As shown in **Table 1** (located at the end of the manuscript), we observed no significant differences between groups regarding age, years of education, the use of hormonal contraception and romantic relationship status. However, groups significantly differed regarding years of education and physical activity. Sleep duration the night before the experimental task was comparable in the three groups.

**Table 1.** Demographic characteristics.

| Variable | Control | BPD + SSRI | BPD – SSRI | p-value |
| --- | --- | --- | --- | --- |
| Body Mass Index (BMI) | 22.24 ± 3.71<br>[18.0–42.0] | 24.14 ± 7.34<br>[17.0–47.0] | 23.85 ± 4.31<br>[18.0–35.0] | 0.229 |
| Age (years) | 25.00 ± 4.40<br>[18.0–35.0] | 22.57 ± 4.36<br>[18.0–33.0] | 25.07 ± 5.18<br>[19.0–35.0] | 0.200 |
| Years of education | 16.98 ± 2.67<br>[11.0–25.0] | 14.21 ± 2.89<br>[11.0–22.0] | 14.63 ± 2.87<br>[11.0–21.0] | < .001 |
| Physically active (yes) | 37 (80.4%) | 4 (28.6%) | 15 (55.6%) | < .001 |
| Contraceptive use (yes) | 15 (32.6%) | 5 (35.7%) | 6 (22.2%) | 0.564 |
| In a romantic relationship (yes) | 32 (69.6%) | 8 (57.1%) | 14 (51.9%) | 0.295 |
| Sleep duration before the experimental task (hours) | 8.17 ± 1.42<br>[5.0–11.0] | 8.97 ± 1.59<br>[6.2–12.3] | 8.34 ± 1.46<br>[4.5–11.0] | 0.202 |

*Note. Continuous variables are reported as mean ± SD [range]; categorical variables represent the percentage of participants answering 'yes'. p-values are based on one-way ANOVA (continuous) or Chi-squared tests (categorical). BPD + SSRI = women with borderline personality disorder taking selective serotonin reuptake inhibitors; BPD – SSRI = women with borderline personality disorder not taking SSRIs.*

Groups significantly differed regarding most clinical symptoms, emotion regulation and attachment styles (**Table 2**, located at the end of the manuscript). Differences were driven by the control group.

**Table 2.**
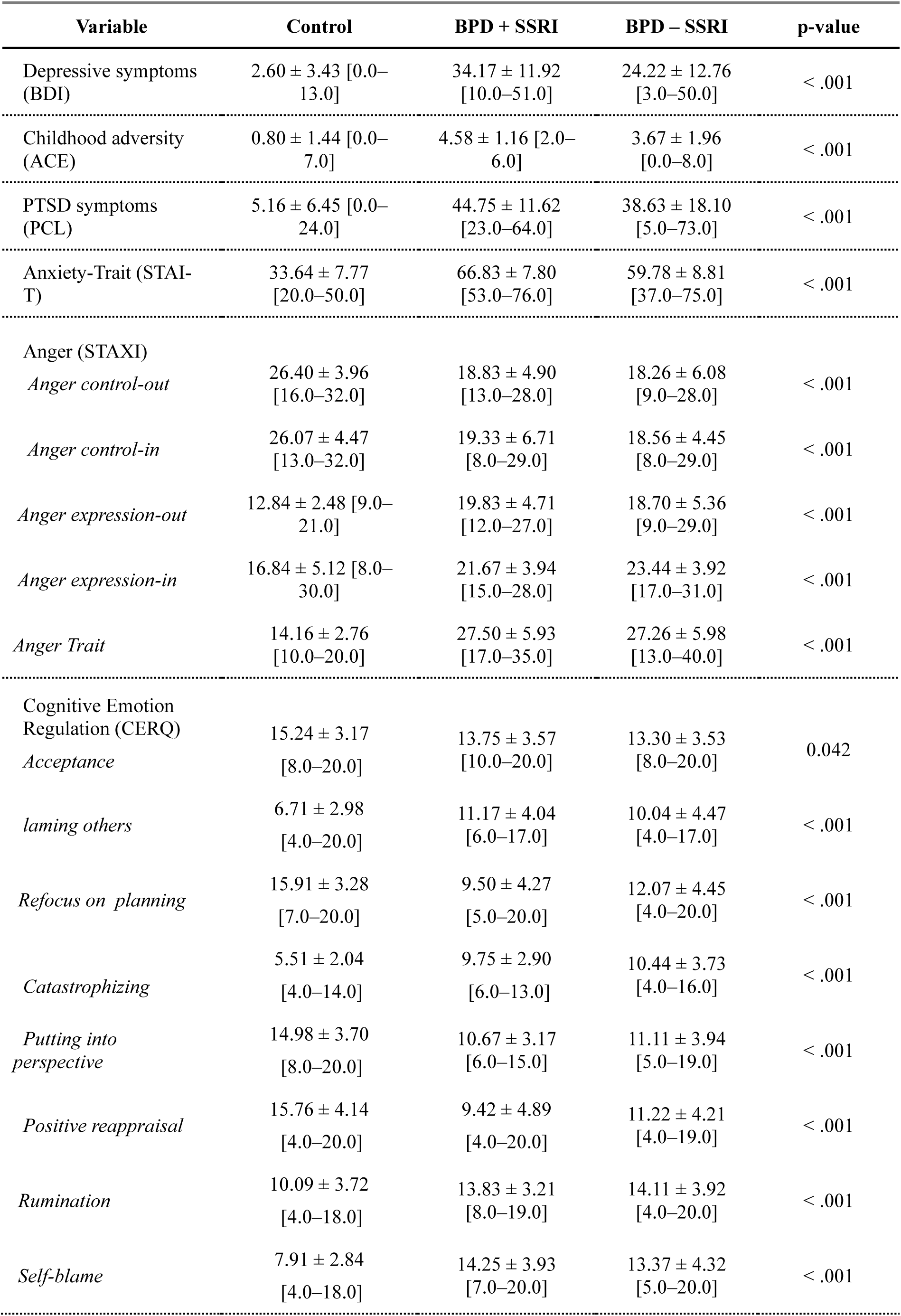
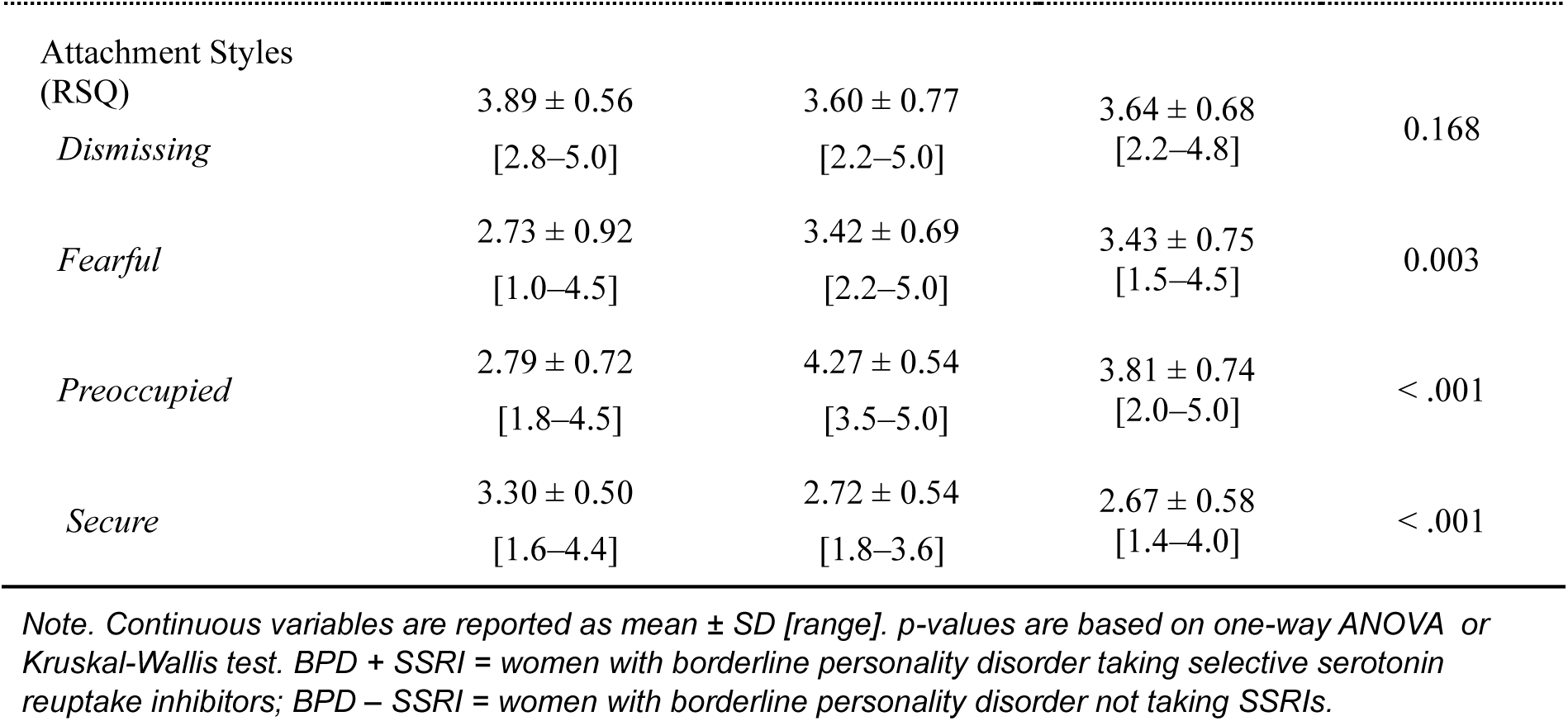
Clinical and symptom scores by group.

### 3.2 Self-reported and hormonal data during the experimental stress task

#### Self-perceived anxiety and anger

Significant main effects of time (F( 2, 168) = 84.06, p < .001) and group ((2, 84) = 57.06, p < .001) were observed for STAI-S scores, but no significant group × time interaction (F(4, 168) = 1.32, p = .263). As for STAXI-state, significant main effect of time (F(2, 168) = 20.35, p < .001) and group F(2, 84) = 10.66, p < .001) were also observed, together with a significant group × time interaction F(4, 168) = 4.13, p = .003, indicating that there was a change over time across goups **(Figure 2)**.

**Figure 2.**
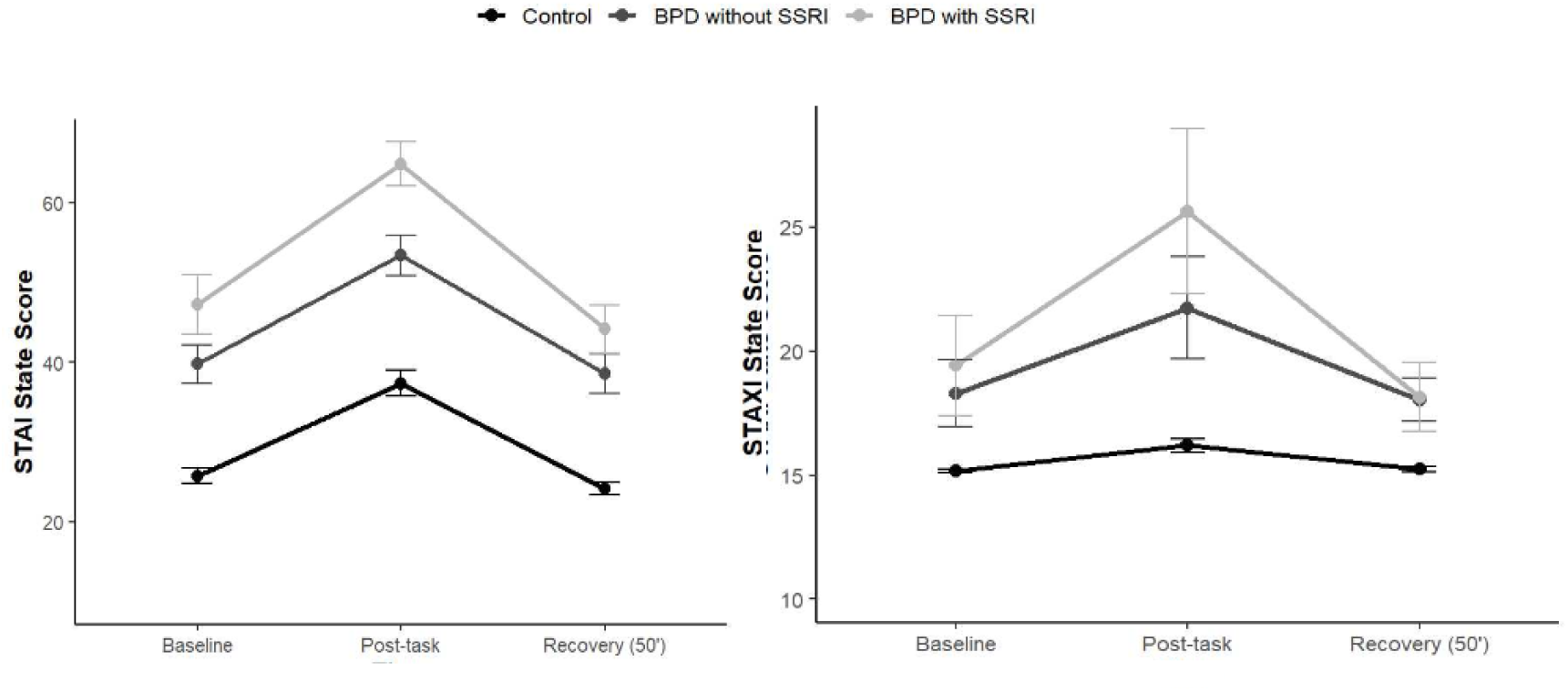
STAI-S and STAXI-S scores during the experimental task.

#### Salivary oxytocin

Linear mixed-effects models applied to OXT values **(Figure 3**) revealed a significant main effect of time, characterized by a specific increase during Anticipation (*B* = 0.06, *t* = 2.24, *p* = .026). There was no significant main effect of group, but a significant time × group interaction, reflcting differential group patterns over time. Specifically, BPD participants not using SSRIs showed a sharper decrease in OXT compared to controls (*B* = −0.12, *t* = −2.60, *p* = .010), whereas BPD participants with SSRI displayed a trajectory comparable to controls.

**Figure 3.**
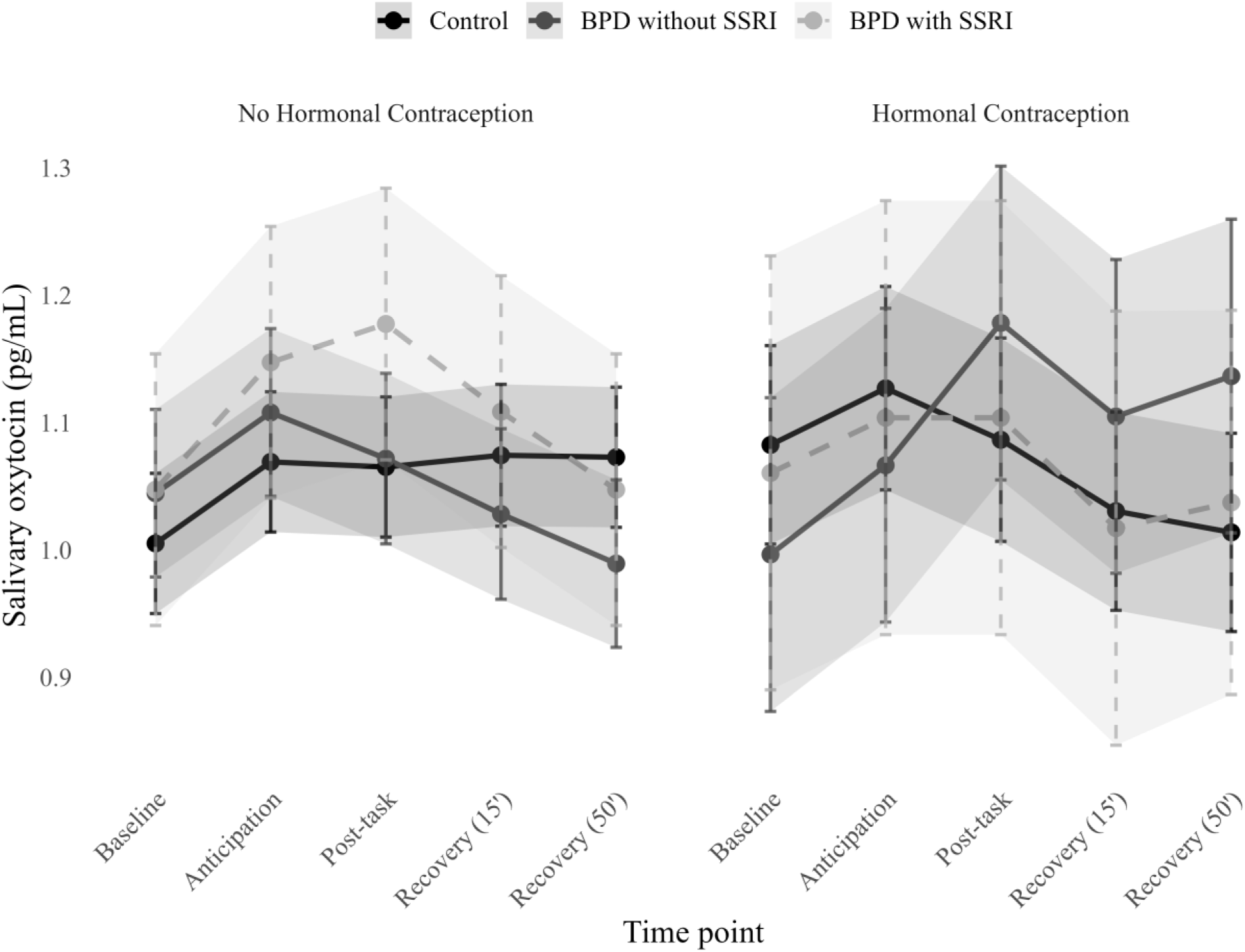
Estimated marginal means of salivary oxytocin in controls across time-points during the TSST for healthy controls, BDP without SSRI and BPD with SSRI; with and without hormonal contraception.

When hormonal contraception was added as a covariate, a significant higher-order interaction with time and group was observed. Among BPD participants not using SSRIs, contraceptive use reversed the OXT pattern seen in non-users, resulting in a progressive increase in OXT at later time points (Post-Task*: B* = 0.21, *t* = 2.25, *p* = .026; Recovery (15’ min): *B* = 0.25, *t* = 2.64, *p* = .009; Recovery (50’ min): *B* = 0.33, *t* = 3.57, *p* < .001).

#### Salivary Cortisol

General linear models applied to CORT values **(Figure 4**) revealed a significant main effect of time. Specifically, significant fluctuations were observed during Anticipation (B = -0.22, z = 2.70, *p* = 0.007), Recovery 15’ min (B = 0.18, z = 2.23, *p* = 0.03) and Recovery 50’ min (B = -0.21, z = -2.55, *p* = 0.02). While there was no significant main effect of group, a significant interaction between time and group emerged (B = -0.32, z = -1.97, *p* = 0.049). Including hormonal contraception as a covariate in the model did not alter this pattern.

**Figure 4.**
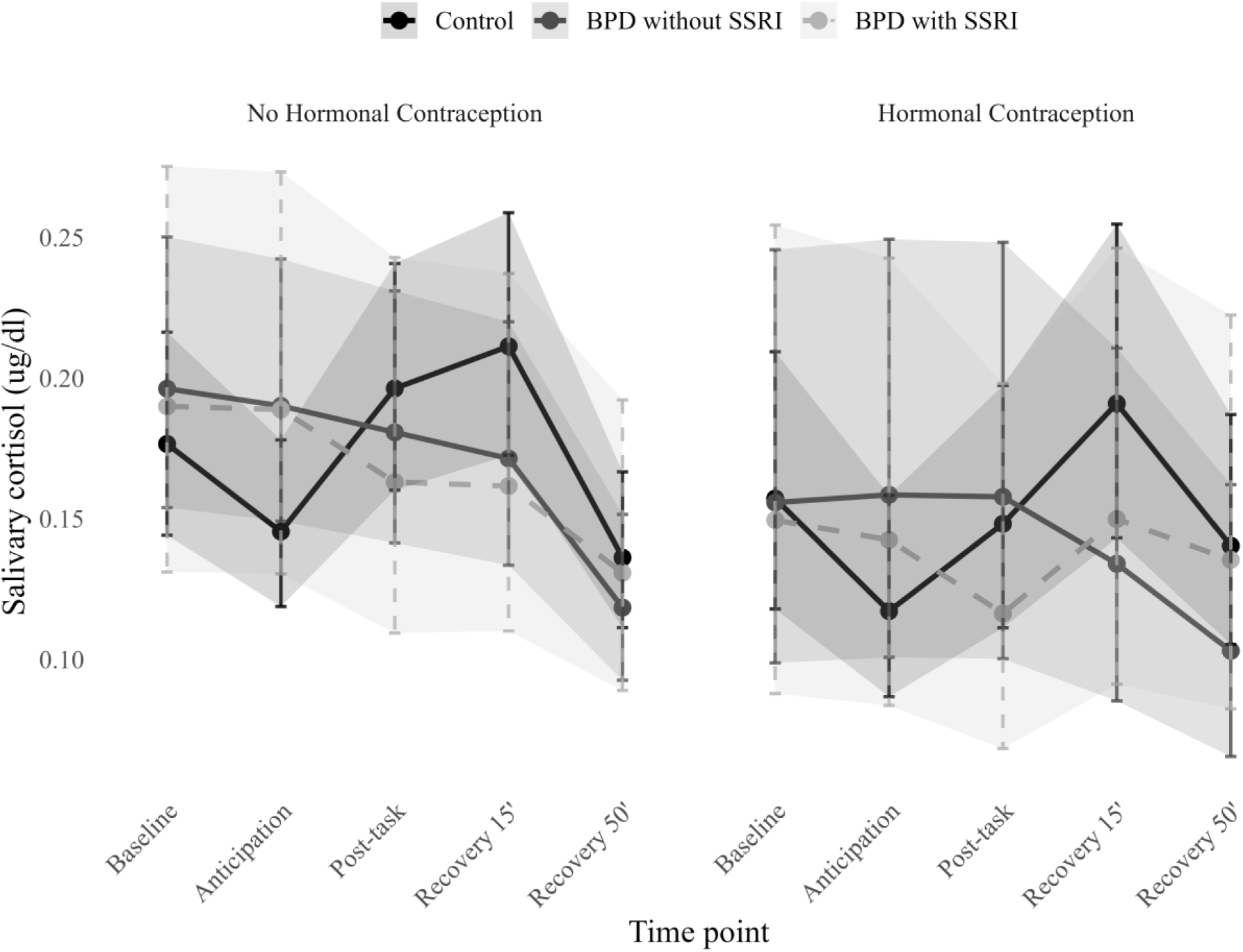
Estimated marginal means of salivary cortisol in controls across time-points during the TSST for healthy controls, BDP without SSRI and BPD with SSRI; with and without hormonal contraception.

#### Interplay between Cortisol and Oxytocin

A significant three-way interaction between oxytocin, group, and time indicated that the association between OXT and CORT **(Figure 5)** varied across groups and time points. Specifically, in BPD participants with SSRI, higher OXT levels were associated with higher CORT concentrations during Recovery 50’ min (*B* = 3.55, *p* = .012), whereas this relationship was weaker and only marginal in BPD participants without SSRI (*B* = 1.96, *p* = .060). No significant oxytocin–cortisol association was observed in the control group.

**Figure 5.**
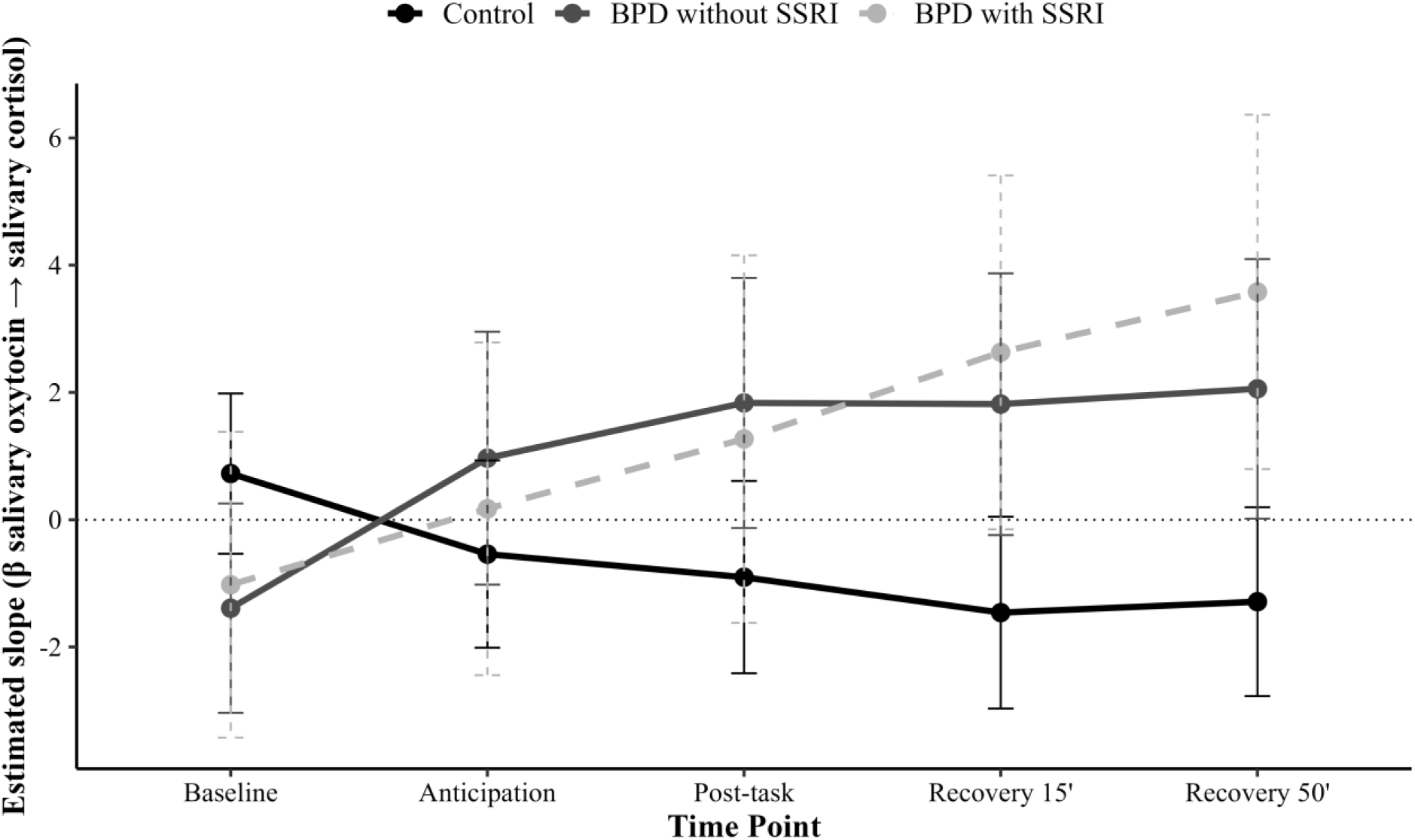
Schematical representation of the temporal dynamics of oxytocin-cortisol interplay.

## 4. Discussion

This experimental study examined OXT and CORT reactivity to a psychosocial stressor in patients with BPD (with and without SSRI treatment) and healthy controls. It further examined the influence of hormonal contraception on these responses and the interplay between OXT and CORT. As expected, the two BPD groups displayed significantly higher subjective (perceived) reactivity to stress compared to controls. Regarding hormonal responses, both OXT and CORT showed significant modulation across time-points, with significantly different patterns between groups over time. For oxytocin in particular, these patterns were modified when inserting hormonal contraception as a covariate.

The higher subjective stress observed in the BPD groups during the task is line with previous studies and compatible with clinical manifestations of BPD (Lieb et al., 2010; Stoffers-Winterling et al., 2012). It confirms that the task was able to induce significant stress and serves as a manipulation check, allowing us to analyse hormonal data occurring under stress conditions. Indeed, differentiating basal and stress-induced secretion is important, as individuals with BPD may maintain relatively apparent normal functioning during periods of stability, with emerging dysregulation when the system is challenged by psychosocial and emotional demands.

Salivary OXT varied significantly across time points in all three groups, characterized by a clear and consistent increase between rest and the anticipation phase (the period after participants had finished preparing their 5-minute speech but had not yet presented it to the examiners). This initial rise was followed by distinct group patterns of post-task activation and recovery. Interestingly, BDP participants not taking SSRI’S displayed a decline in OXT concentrations during and after the stress task, while BPD participants under SSRIs displayed trajectories comparable to controls. This pattern is consistent with earlier reports of attenuated OXT responsiveness to psychosocial stress in BPD patients (Jobst et al., 2014; Aboulafia-Brakha et al., 2023) and evidence of SSRI modulation of OXT production and availability on the central nervous system (Brune, 2015, Bommann et al. 2017). While previous studies did not differentiate patients based on pharmacological treatment, our study adds this important distinction. However, it is relevant to point out that although SSRIs appear to normalize the hormonal stress response in patients with BPD in a laboratory setting, clinical evidence indicates no significant effect of SSRIs on the core symptoms of the disorder (Lieb et al., 2010; Stoffers-Winterling et al., 2012).

Further, when hormonal contraception was included as a covariate in the OXT reactivity model, we observed a change in the response pattern in the BPD group without SSRIs, resulting in OXT secretion during the post-task and recovery phases that mirrored the control group and the BPD SSRI group without hormonal contraception. This influence suggests a synergistic modulation between sex hormones and OXT regulation, consistent with data showing higher OXT levels in women using estrogen-containing contraceptives (Engert et al., 2016; de Jong et al., 2015). Indeed, estrogen seems to enhance OXT synthesis and receptor sensitivity (Thomas et al., 2019), which may account for the observed recovery-phase increase. However, the lack of a similar effect in BPD participants using SSRIs suggests a potential convergence on shared neuroendocrine pathways. Because both SSRIs and estrogen modulate oxytocin release through 5-HT1A receptor signaling, these pathways may be sufficiently activated or desensitized by either molecule, leading to a state of mutual interference where the additional influence of hormonal contraception is no longer observable (Brüne, 2015; Bomann et al., 2017).

Regarding CORT, distinct group patterns were observed across time, with particular hyporeactivity in the BPD group without SSRIs. This result replicates earlier findings of blunted HPA-axis activation in BPD (Deckers et al., 2014; Bourvis et al., 2017), suggesting maladaptive physiological regulation in response to interpersonal stressors. However, hormonal contraception did not affect this pattern, suggesting that glucocorticoid pathways are less influenced than OXT pathways by sex hormones (Thomas et al., 2019; de Jong et al., 2015, Quintana et al. 2024). Further, the CORT peak response was observed 15 min after the end of the task in healthy controls, which is also consistent with previous reporting and the observation that OXT peaks earlier than cortisol during stress (Engert et al., 2016; Bernhard et al., 2018; Alley et al., 2019, (Peled-Avron et al., 2020). Indeed previous studies suggest that OXT has a moderating role on CORT reactivity Heinrichs & Domes, 2008). In line with this we found that OXT affected CORT pathways across time as shown by a significant interaction between OXT, group and time for the CORT response. Further analyses would be needed to explore the temporal relationship of this interplay.

Several limitations should be considered when interpreting these findings. First, the absence of a clinical control group (e.g., patients with depression or anxiety disorders without BPD) limits our ability to determine whether the observed neuroendocrine patterns are specific to BPD or reflective of general transdiagnostic distress. Second, we did not randomly assign medication or contraception status, which prevents causal conclusions regarding the specific effects of SSRIs or sex hormones. It is possible that patients prescribed pharmacological treatments differ in clinical severity from those who are not (indication bias). Finally, while salivary OXT is a widely accepted non-invasive measure, questions regarding the precise correlation between peripheral concentrations and central oxytocinergic activity remain a subject of methodological debate. Despite these constraints, the study provides a controlled examination of how common treatments interact with stress physiology in a standardized setting (Heinrichs et al., 2003; Simeon et al., 2011).

## Conclusion

In conclusion, this study confirms that patients with BPD experience heightened subjective distress in response to psychosocial pressure, a finding that aligns with the disorder’s core emotional dysregulation. Crucially, our results highlight the importance of analyzing neuroendocrine markers under stress conditions, as specific dysregulations in Oxytocin and Cortisol output were only revealed during the dynamic challenge of the task. Furthermore, these biological patterns appear sensitive to pharmacological influence; both SSRI treatment and hormonal contraception significantly modulated—and in some cases ‘normalized’—the hormonal stress response. However, it is essential to interpret this biological plasticity with caution. While SSRIs may stabilize neuroendocrine reactivity in a laboratory setting, this does not necessarily translate to clinical improvement, consistent with guidelines indicating limited efficacy of SSRIs for treating BPD core symptoms. Future research must bridge this gap, clarifying how these sensitive biological markers relate to long-term therapeutic outcomes.

## Data Availability

All data produced in the present study are available upon reasonable request to the authors

